# District level correlates of COVID-19 pandemic in India

**DOI:** 10.1101/2020.10.08.20208447

**Authors:** Vandana Tamrakar, Ankita Srivastava, Mukesh C. Parmar, Sudheer Kumar Shukla, Shewli Shabnam, Bandita Boro, Apala Saha, Benjamin Debbarma, Nandita Saikia

**Affiliations:** Centre for the Study of Regional Development, Jawaharlal Nehru University, New Delhi, India; Banaras Hindu University, Varanasi, India; Bidhannagar College, Kolkata, India

**Keywords:** COVID-19, socio-economic, co-morbidity, geographical, hot-cold spot, districts, India

## Abstract

**Background:** The number of patients with coronavirus infection (COVID-19) has amplified in India. Understanding the district level correlates of the COVID-19 infection ratio (IR) is therefore essential for formulating policies and intervention.

**Objectives:** The present study examines the association between socio-economic and demographic characteristics of India’s population and the COVID-19 infection ratio at district level…

**Data and Methods:** Using crowdsourced data on the COVID-19 prevalence rate, we analyzed state and district level variation in India from March 14 to July 31 2020. We identified hotspot and cold spot districts for COVID-19 cases and infection ratio. We have also carried out a regression analysis to highlight the district level demographic, socio-economic, infrastructure, and health-related correlates of the COVID-19 infection ratio.

**Results:** The results showed that the IR is 42.38 per one hundred thousand population in India. The highest IR was observed in Andhra Pradesh (145.0), followed by Maharashtra (123.6), and was the lowest in Chhattisgarh (10.1). About 80 per cent of infected cases, and 90 per cent of deaths were observed in nine Indian states (Tamil Nadu, Andhra Pradesh, Telangana, Karnataka, Maharashtra, Delhi, Uttar Pradesh, West Bengal, and Gujarat). Moreover, we observed COVID-19 cold-spots in central, northern, western, and north-eastern regions of India. Out of 736 districts, six metropolitan cities (Mumbai, Chennai, Thane, Pune, Bengaluru, and Hyderabad) emerged as the major hotspots in India, containing around 30 per cent of confirmed total COVID-19 cases in the country. Simultaneously, parts of the Konkan coast in Maharashtra, parts of Delhi, the southern part of Tamil Nadu, the northern part of Jammu & Kashmir were identified as hotspots of COVID-19 infection. Moran’s-I value of 0.333showed a positive spatial clusteringlevel in the COVID-19 IR case over neighboring districts. Our regression analysis found that district-level population density (β: 0.05, CI:004-0.06), the percent of urban population (β:3.08, CI: 1.05-5.11), percent of Scheduled Caste Population (β: 3.92, CI: 0.12-7.72),and district-level testing ratio (β: 0.03, CI: 0.01-0.04) are positively associated with the prevalence of COVID-19.

**Conclusion:** COVID-19 cases were heavily concentrated in 9 states of India. Several demographic, socio-economic, and health-related variables are correlated with COVID-19 prevalence rate. However, after adjusting the role of socio-economic and health-related factors, the COVID-19 infection rate was found to be more rampant in districts with a higher population density, a higher percentage of the urban population, and a higher percentage of deprived castes and with a higher level of testing ratio. The identified hotspots and correlates in this study give crucial information for policy discourse.

## Introduction

With more than16,96,962 confirmed cases on July 31,2020, India ranked third globally in terms of the total number of infected patients of COVID-19(1). The rate of spread of the disease was slow in the initial three months of the first outbreak in Kerala in January 2020, possibly because of the early nationwide lockdown(2–4); widespread coverage about the pandemic in print, electronic and social media(5), and targeted efforts by the union and state governments on quarantine facilities and travel protocols (6,7). With the demarcation of local containment zones, these definitive measures significantly reduced the doubling time **(S1Fig**.), although there is no sign of stalling in the infection rates. There is a rapid increase in the number of confirmed cases of COVID-19 in many districts. India has been recording over 50,000 new cases every day since July 25, 2020. It took 59 days (14 March-18 May) to increase from about 1,000 cases to more than 1,00,000 cases (Figure 1), whereas it took only 15 days (May 19-June 2) to add 100,000 new COVID-19 patients. Afterward, India is adding one hundred thousand new confirmed cases every two days.

**Fig 1.**
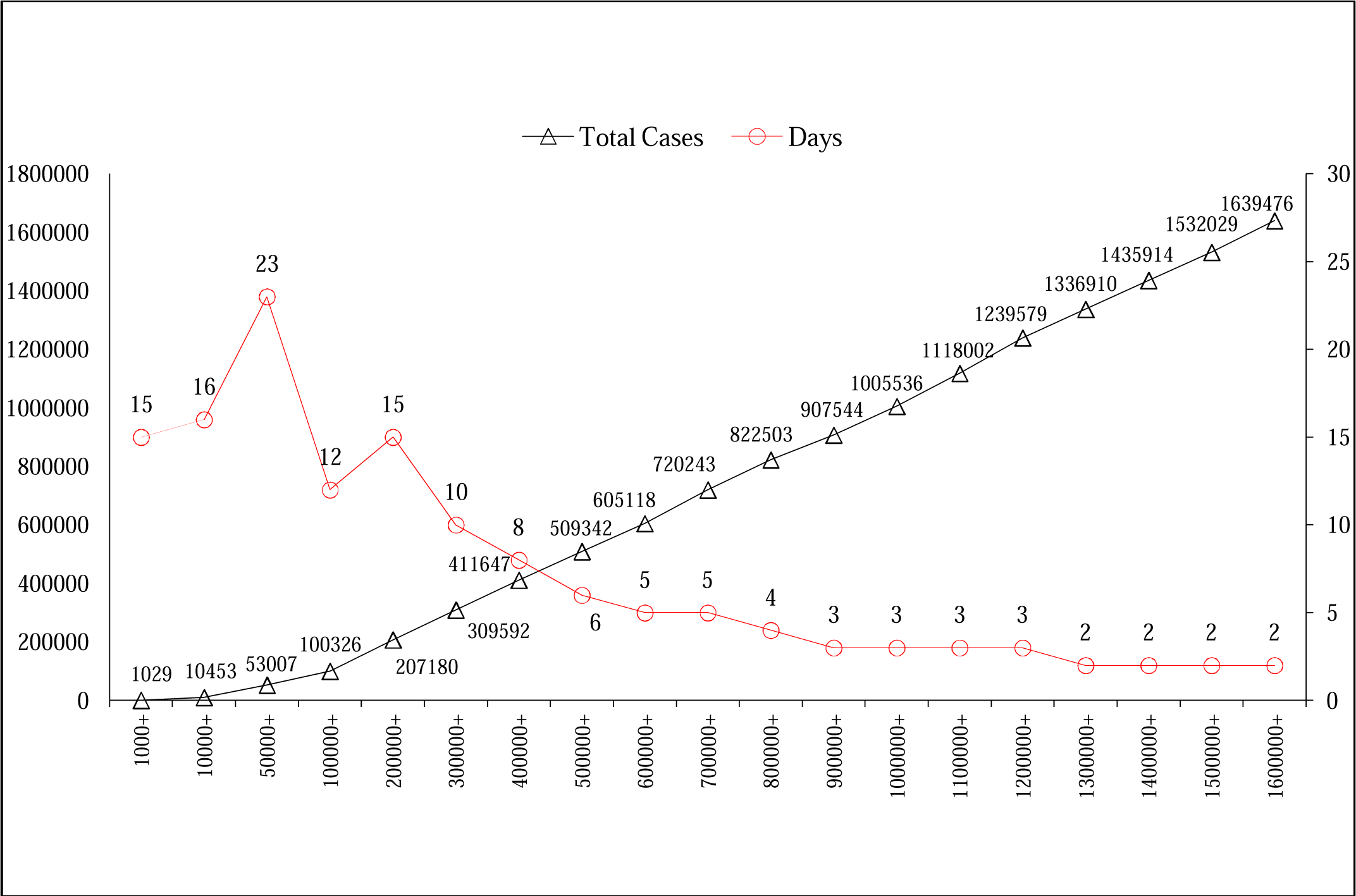
Trend in new cases by number of days in India, March 14- July 30, 2020. Trend of number of total confirmed cases by no. of days in India 14 March-July 30, 2020 Source: Author’s calculations

**Fig 2.**
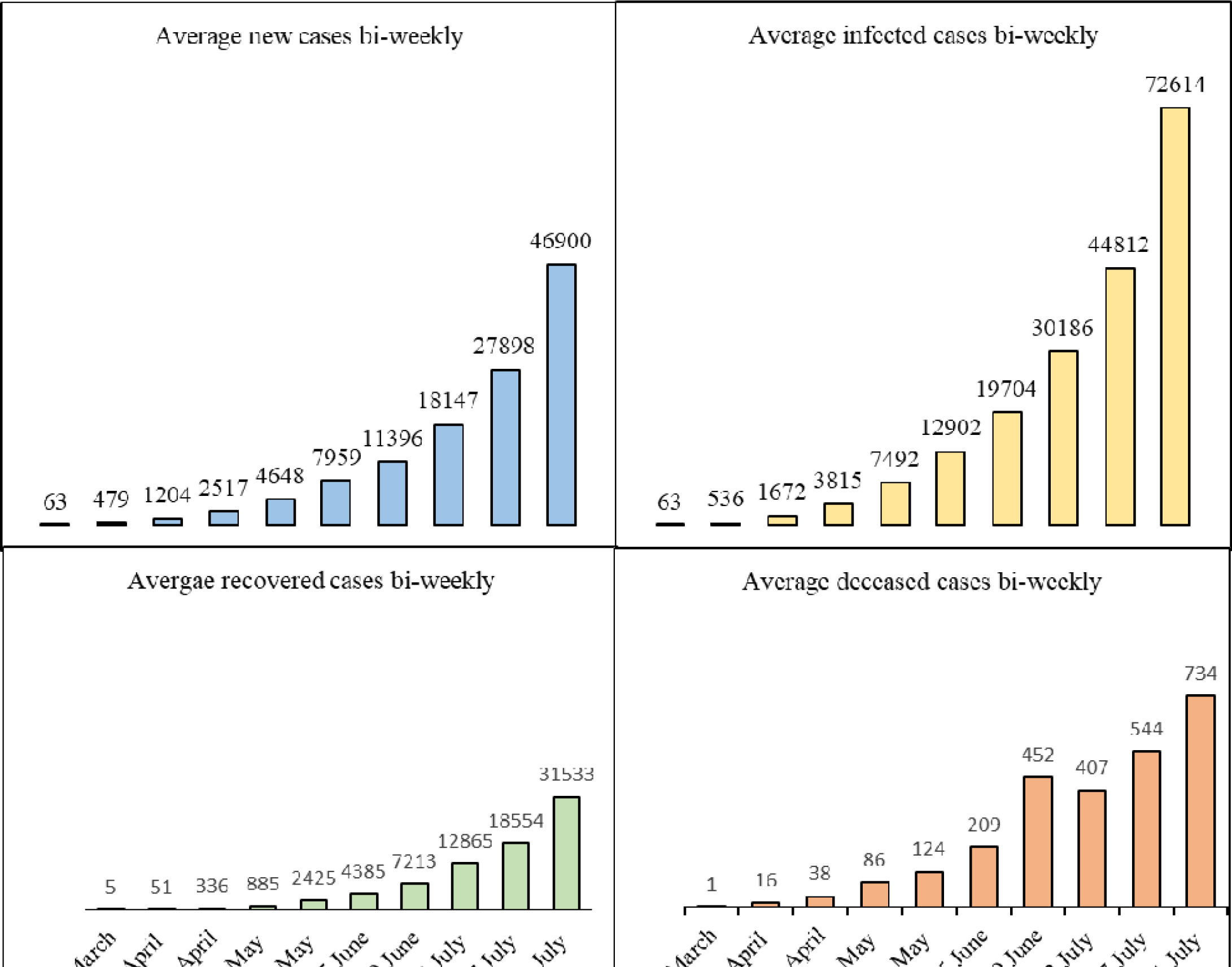
Bi-weekly average new, infected, recovered, deceased cases in India (March 14 - July 31, 2020) Source: Author’s calculations

**Fig 3.**
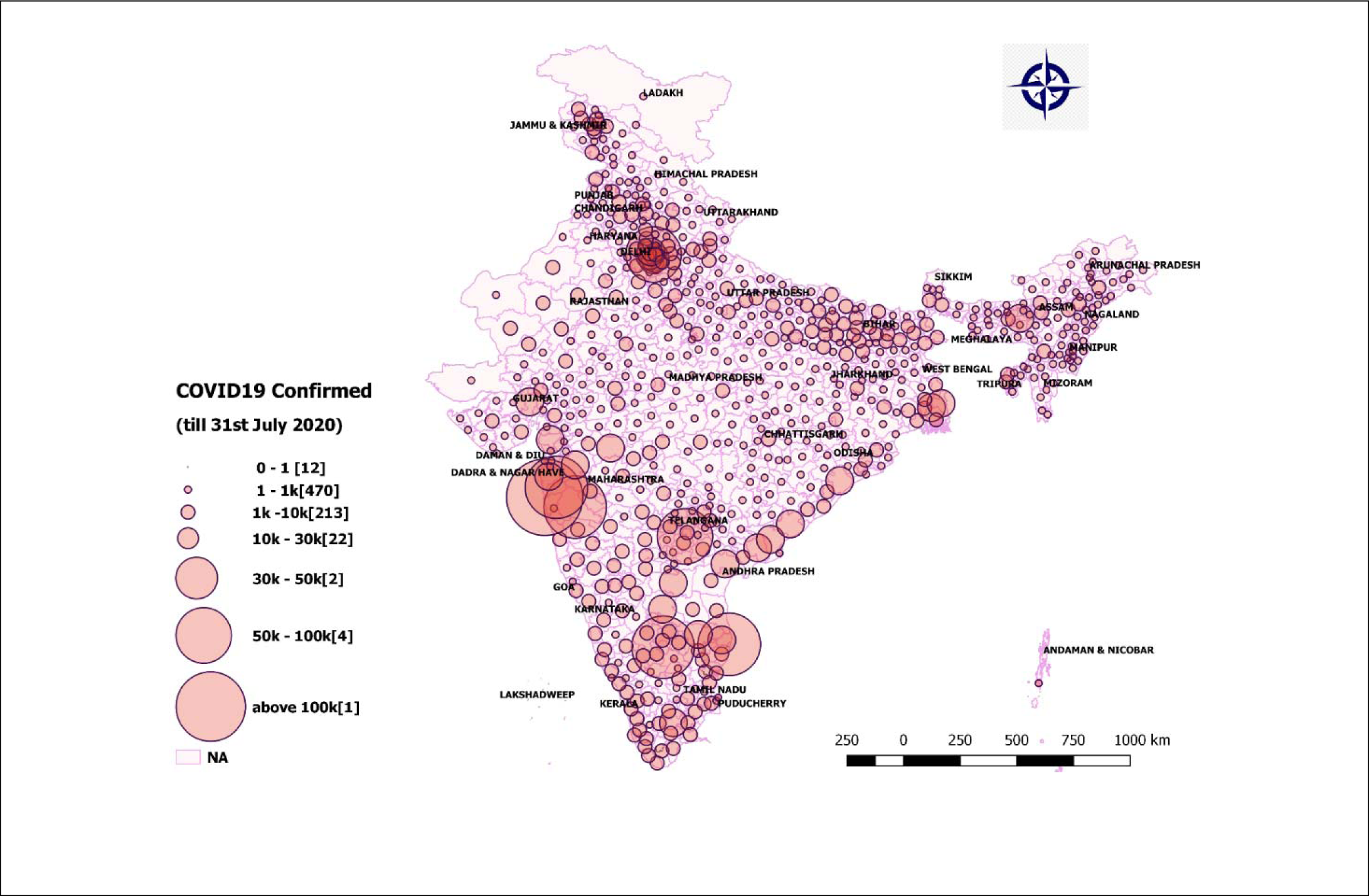
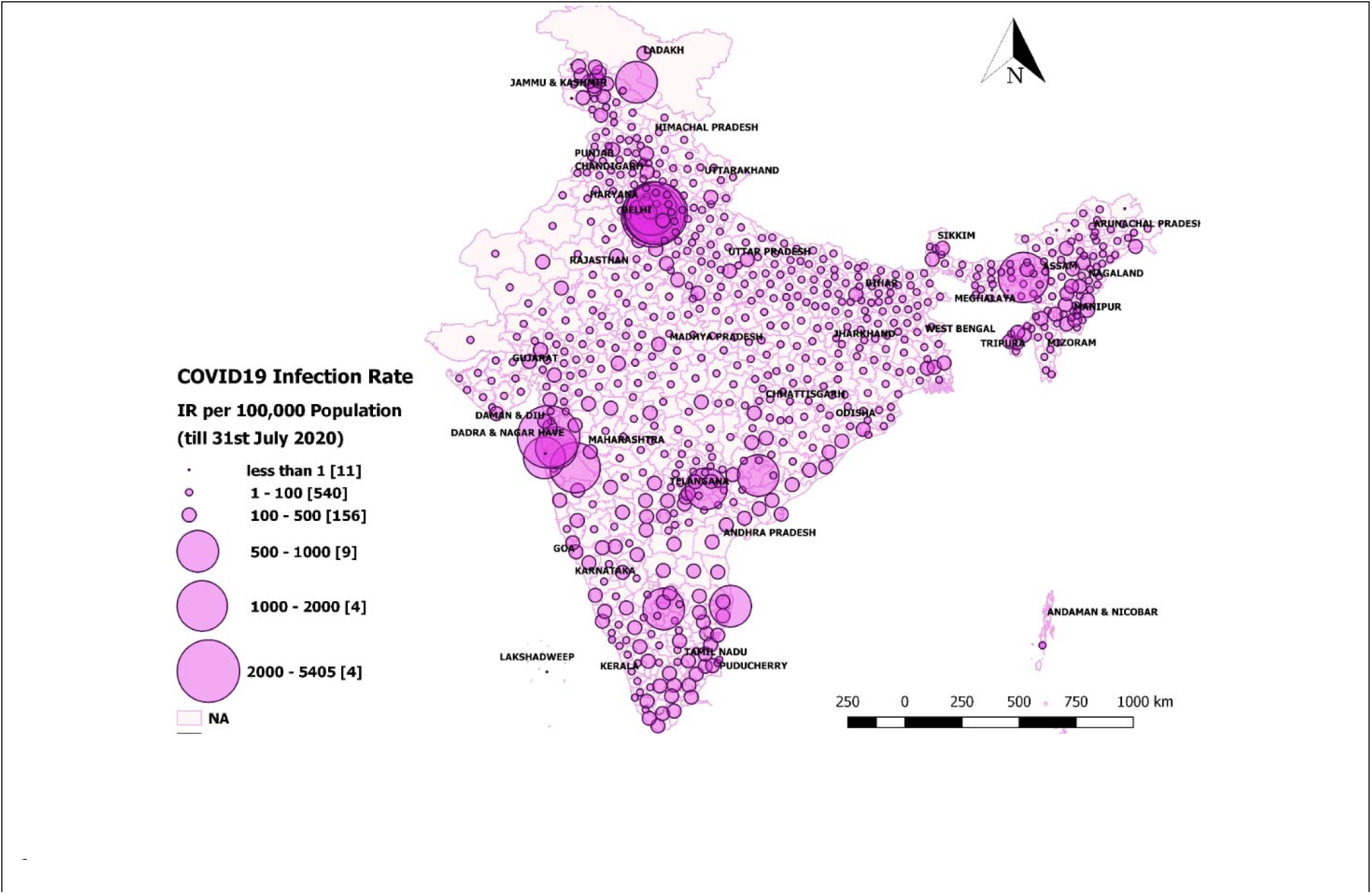
District-level variations in COVID-19 on July 31, India 2020. **Panel A:** Number of positive (absolute values) COVID-19 cases in districts as of July 31, 2020 **Panel B:** District level infection ratio (IR), defined as the number of confirmed cases per 100,000 population by July 31, 2020 **Source:** Author’s calculation

**Fig 4.**
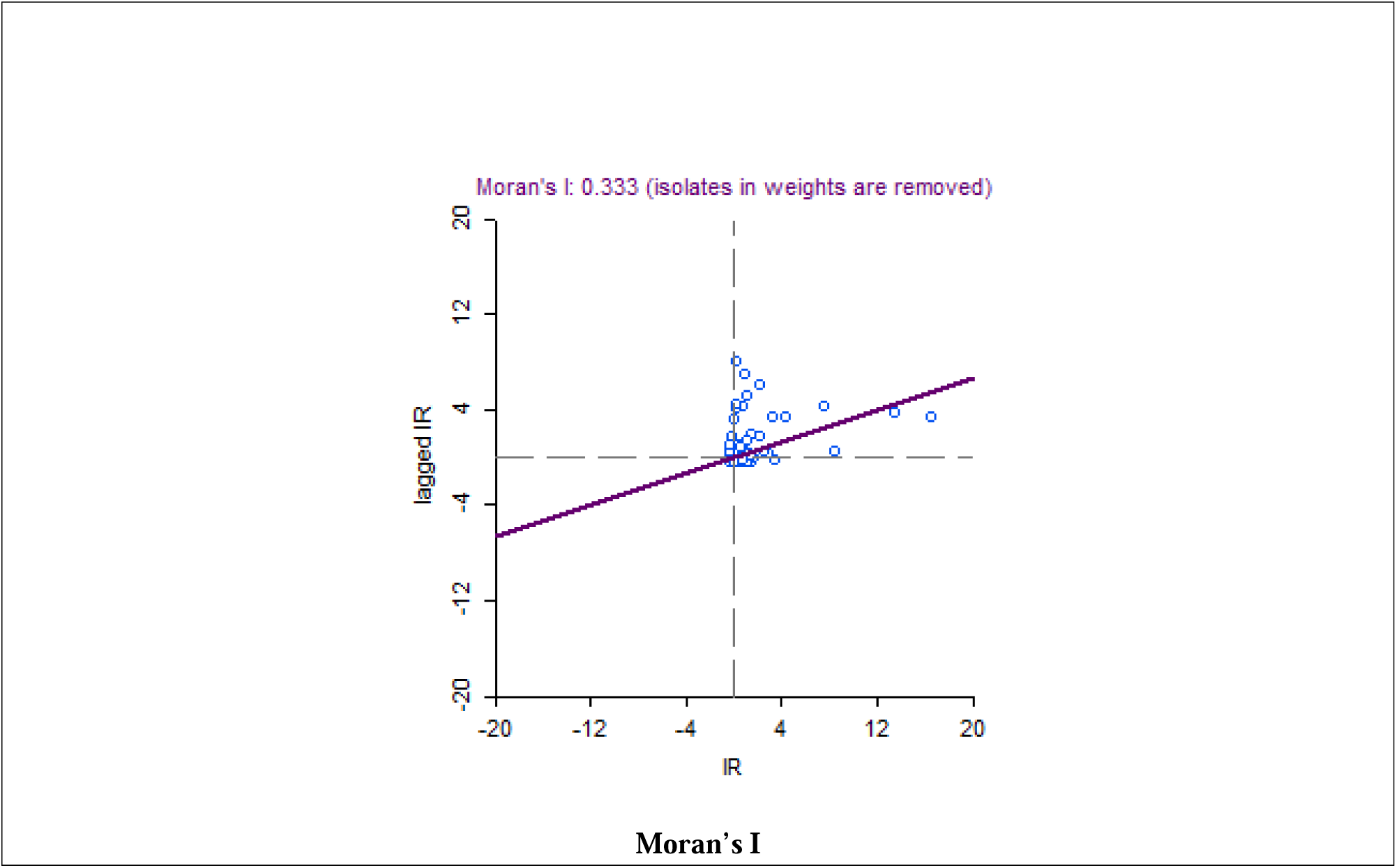
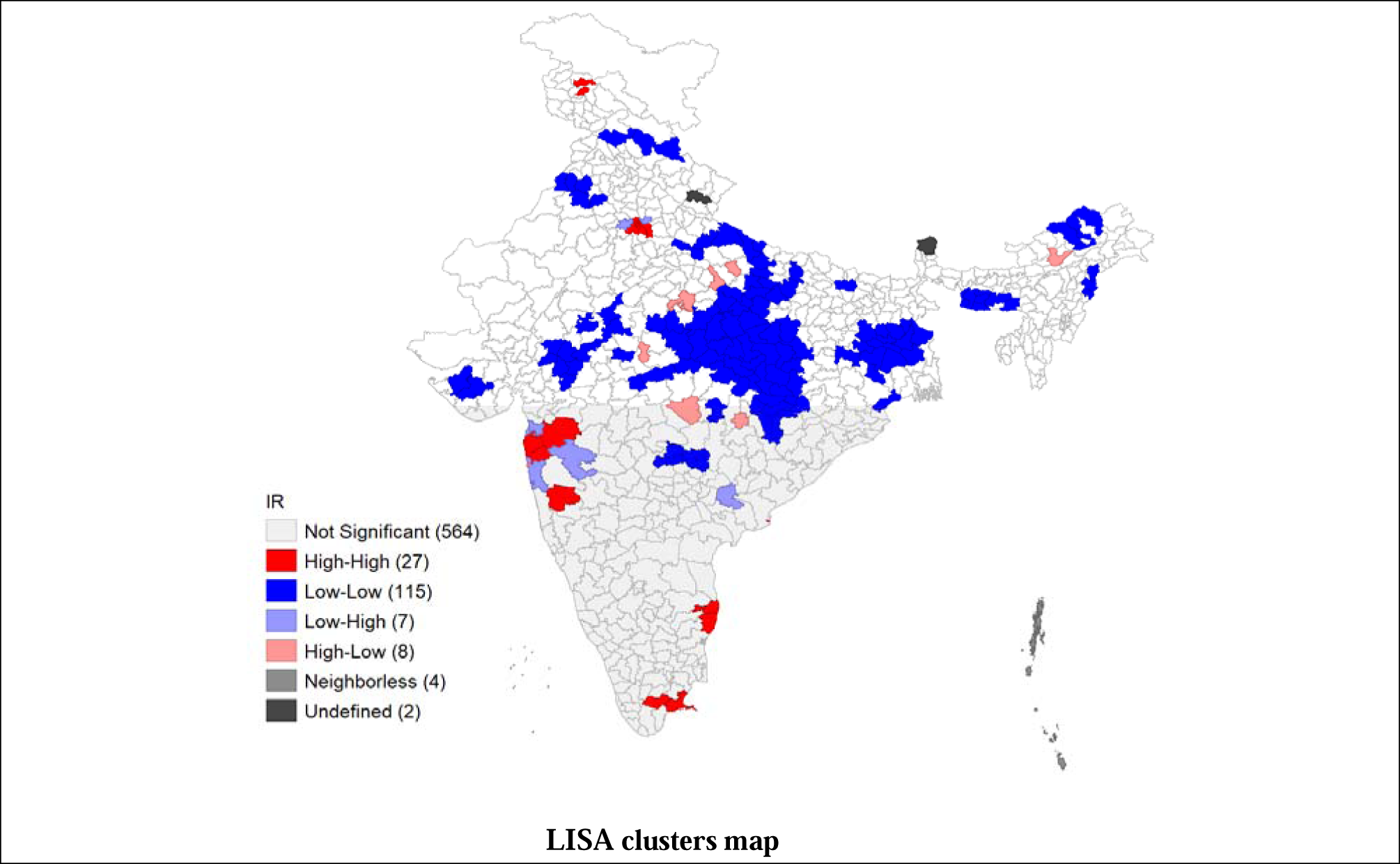
Moran’s I and LISA cluster maps for district’s infection ratio of COVID-19 in India till July 31 2020. **Source:** Author’s calculations

Despite such a fast spread of COVID-19, India has a fairly high recovery rate and the lowest fatality rate globally(8). Despite India’s advantage of having a young age structure less susceptible to COVID-19 related deaths(9). India may have to undergo a higher burden of disease shortly due to other demographic factors (10) such as the enormous population size, high population density, higher percentage of people living in poverty, lower levels of per capita public health infrastructure, and a high prevalence of co-morbid situations.

Like any other health and demographic indicator, COVID-19 infection varies widely among the different states of the country (11,12). However, the geographical pattern of the COVID-19 infection rate’ does not coincide with the patterns of demographic and health indicators such as the under-five mortality rate or nutritional status. COVID-19 has been spreading rapidly in the urban areas, especially in states with megacities with densely populated urban slums like Delhi, Maharashtra, Tamil Nadu, and West Bengal. The sudden surge of return labour migration to the states of origin (due to COVID-19 related national lockdown), state-level health care system, adherence to physical distancing measures, and local government management are other potential community-level factors affecting geographical variations in the spread of COVID-19 in India. Some recent studies have computed composite indices to rank the districts in terms of their COVID-19 vulnerabilities using demographic information and infrastructure characteristics(13–15). While such analyses are useful for district-level planning and prioritization they are based on the assumption that vulnerability will decrease as -the districts’ socio-economic indicators improve. However, such an inverse relationship may not be applicable in the rare COVID-19 context; for instance, a higher percentage of urban population may indicate a higher socio-economic status of the district population in a non-COVID situation but maybe positively correlated with the spread of COVID-19. COVID-19 is more prevalent in cities and towns than in rural areas or hilly regions(16).Therefore, it is imperative to unfold the empirical relationship patterns between the district’s socio-economic and infrastructural characteristics and the COVID-19 infection ratio. To the best of our knowledge, no such previous study has been conducted on COVID-19 in India. This study examined the district level socio-economic and demographic correlations of COVID-19 infection ratio in India. Identification of such correlates is crucial for framing health policy and appropriate intervention.

## Data and Methods

We used crowdsourced district-level data on COVID-19 available in the public domain until July 31, 2020, accessed from the COVID-19India dashboard(17). It is an application programming interface (API) to monitor the COVID-19 cases at national, state, and district levels. The data compiled in this web portal is based on state bulletins and official handles. The details of the data are available on the website. This portal data is consistent with the data provided by the Ministry of Health and Family Welfare, Government of India (https://www.mohfw.gov.in/) (18).

For explanatory variables, we utilized data from the National Family Health Survey of India 2015-16 (NFHS-4), a cross-sectional survey of 601,599 households, and 2.87 million individuals from all 29 states and 7 union territories (19). The survey collected data on various socio-economic, demographic, health, and family planning indicators and anthropometry and biomarkers’ measures related to anemia, hypertension, and diabetes. The NFHS-4 is the most recent source of such biomarker-based data at the district level in India. We also used some socio-economic and demographic variables from the Census of India(20).

### Outcome variable

For all the 640 districts in the thirty-five states and eight union territories of India, we defined the outcome variable, COVID-19 Infection Ratio (IR), as the number of confirmed cases in a given district per 100,000 population. For the district-level population for the year 2020, we projected the district population using an exponential growth rate from the census 2001 and 2011.

The infection of ratio was calculated as:

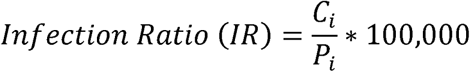

Where,

*C*_*i*_ = the number of confirmed cases in i^th^ district and *P*_*i*_ = total projected population in the i^th^ district on July 31,2020.

### District level correlates

Based on previous literature(21–24), we considered a set of 26 variables at district level, viz., i) Demographic variables: percentage of population aged 60 and above, percentage of women whose husbands are away for the last 6 months, population density, ii)Socio-economic variables: percentage of the literate population, percentage of Scheduled Castes (SC) population, percentage of Scheduled Tribes (ST) population, percentage of Hindu population, percentage of urban population, average number of persons that sleep in one room; iii) Infrastructure variables: percentage of households with availability of soap, percentage of households with water and toilet facility within the premise; iv) Health-related variables: percentage of women with Diabetes (Glucose>140mg),percentage of women (among age18+) with Cancer, percentage of 18+aged women consuming tobacco, testing ratio per one hundred thousand population, under-five mortality rate, percentage of institutional births, percentage of full immunization among children aged 23-36 months, percentage of women aged 18 and above reporting anemia and, percentage of children with stunting and wasting.

### Statistical Analysis

We performed a bi-weekly trend analysis of COVID-19 cases in India. To examine the district level correlates of the outcome variable, we carried out a linear regression analysis at the district level. Two separate district-level regression models were fitted. Model 1 presents the independent variable’s unadjusted effect without controlling the effect of any other independent variable. Model 2 shows the adjusted results of the independent variables on the dependent variable. We did all the analyses in the statistical package Stata14.1. We tested for the possible multicollinearity among the independent variables before fitting them to the regression model.

### Spatial Analysis

We generated descriptive maps of 727 districts in the software package QGIS and later exported the shapefiles to GeoDa software to perform spatial analysis. Using the first-order ‘Queen’s contiguity matrix as the weight, we estimated Moran’s I and univariate Local Indicators of Spatial Association (LISA). ‘Moran’s I” is the Pearson coefficient measure of spatial autocorrelation, which measures the degree to which data points are similar or dissimilar to their spatial neighbours (29). The LISA cluster map yields four types of geographical clustering of the interest variable (30).

Here, “high-high” refers to the regions with above-average infection ratio and sharing the boundaries with neighbouring areas with above-average infection ratio values. On the other hand, “high-low” indicates regions with below-average value and the surrounding areas with above-average infection ratio. The “high-high” are also referred to as *hot spots*, whereas the “low-low” referred to as *cold spots*.

## Results

India has been reporting new cases of the coronavirus (COVID-19) every day since March 14, 2020. India reported over 1696 thousand confirmed cases as of July 31, 2020. Out of these, around 1095 thousand patients have recovered, while 36,565 cases were fatal (17). **S2 Fig**. and **S1 Table** presents the national bi-weekly (14 days) national pattern of new confirmed, infected, recovered, and deceased COVID-19 cases in India. In India, the average bi-weekly new confirmed cases rose 744 times (from 63 to 46,900), the average recovered cases increased 6307 times (from 5 to 31,533), the average infected cases increased 1152 times (from 63 to 72614). The average deceased cases increased 734 times (from 1 to 734) between the 1^st^ and the 10^th^ bi-weekly phases, as given in Table A1. The COVID-19 cases have amplified in each of the Indian states until July 31, 2020. Therefore, the five states of Maharashtra, Andhra Pradesh, Tamil Nadu, Karnataka, and Telangana, have accounted for more than half of the country’s total cases. The interesting fact is that 80 per cent new cases, 79 per cent recovered cases, 80 per cent infected patients, and 90 per cent deaths are from only nine states (Maharashtra, Delhi, Tamil Nadu, Karnataka, Andhra Pradesh, Telangana, West Bengal, Gujarat, and Uttar Pradesh) in India. In the **S2 Table**, we showed the state-wise Infection Rate (IR) per one hundred thousand population till July 31, 2020. India showed that the IR is 42.38 per 100 hundred thousand people. The highest IR was observed in Andhra Pradesh (145.0), followed by Maharashtra (123.59) and lowest in Chhattisgarh (10.12). However, the states like Delhi (54.03), and Southern states Tamil Nadu (76.58), Telangana (45.13), Karnataka (109.43), and Jammu Kashmir (58.81) showed IR above the national average. Only two Union Territories, viz., Lakshadweep, and Daman & Diu, experienced zero cases during the study period.

### District Pattern

**S3 Fig**.in **Panel A** shows the district-level variations in COVID-19. The size of the bubble in the figure indicates the number of positive COVID-19 cases in districts on July 31, 2020. The larger the size of the bubble, the higher is the number of positive cases. Of the 736 districts, five urban districts contain about 28 per cent of the confirmed cases (Mumbai, 7.02 per cent; Chennai, 6.13 per cent; Thane, 5.73 per cent; Pune, 5.48 per cent and Bengaluru, 3.41 per cent). About 11 per cent of the confirmed cases belongs to another seven majorly urban districts (Hyderabad, Central Delhi, Ahmedabad, South East Delhi, Kolkata, West Delhi, and East Godavari,) with at least 20,000 confirmed cases. About85 per cent (625 districts) have at least 100 confirmed cases each, and about 36 per cent (268 districts) have at least one positive confirmed case of COVID-19. About two per cent (12 districts) reported at least one positive case of COVID-19 until July 31, 2020.

**S3 Fig**. in **Panel B** presents the district level infection ratio (IR), defined as the number of confirmed cases per 100,000 population by July 31, 2020.The results show that the top four worst affected districts (Central Delhi, New Delhi, Palghar, Shahdara) in India have at least 2,000 (per 100,000 population) IR. Four districts (South East Delhi, Kamrup Metropolitan, North Delhi, and Pune) have IR ranging between 1000 to 2000 per one hundred thousand population. Nine districts have an infection rate of 500 to 1000 per one hundred thousand people. These are Chennai, West Delhi, Thane, Gurugram, Bengaluru Urban, Mumbai, Jangaon, Faridabad, Papum Pare).

**S4 Fig**. presented the Moran’s I and LISA cluster maps for the district infection ratio of COVID-19 in India. The ‘Moran’s I value of 0.333 represents a positive spatial clustering level in the COVID-19 infection rate over neighbouring districts. Hotspots of COVID-19 were observed in the parts of Konkan coast of Maharashtra (Palghar, Mumbai, Thane, Nashik, and Satara); the southern part from Tamil Nadu (Chennai, Chengalpattu, Thiruvallur, Virudhunagar, and Ramanathapuram); parts of Delhi; the northern part of Jammu & Kashmir (Ganderbal and Pulwama) whereas cold-spots were observed in central, north-western and north-eastern regions of India.

**Table 1** shows the descriptive statistics of the outcome variable (Infection Ratio) and the 26 selected exposure variables for the 640 districts. The average infection ratio is 108.40, and the values vary between 0 and 5918.43 per one hundred thousand population. The IR is zero in the two districts from Arunachal Pradesh (Dibang valley, KrumungKumey) and another two districts from the Union Territories (Lakshadweep and Nicobar). All district-level socio-economic variables differ substantially among districts. For example, the mean percent of the old age population (60 and above) is 8.33, and it varies between 2.46 per cent to 17.82 per cent among the districts of India. The percent of women whose husbands are away for the last 6 months, a proxy indicator of district-level outmigration, ranged substantially between 1.36 per cent to 33.60 per cent. Population density (number of persons per square kilometer) varies from 1 in Dibang Valley (Arunachal Pradesh) to 36155in northeast Delhi.

**Table 1.**
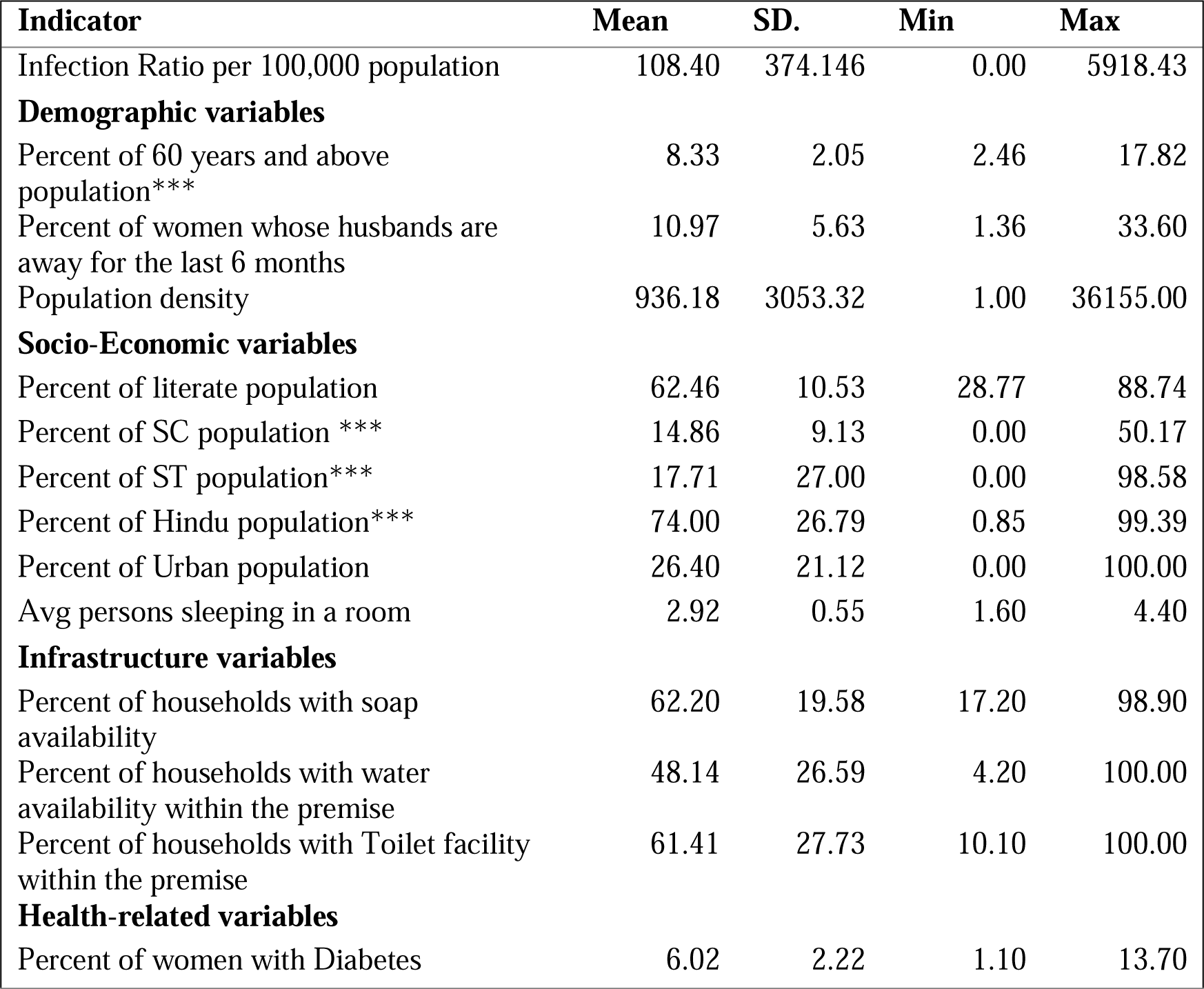
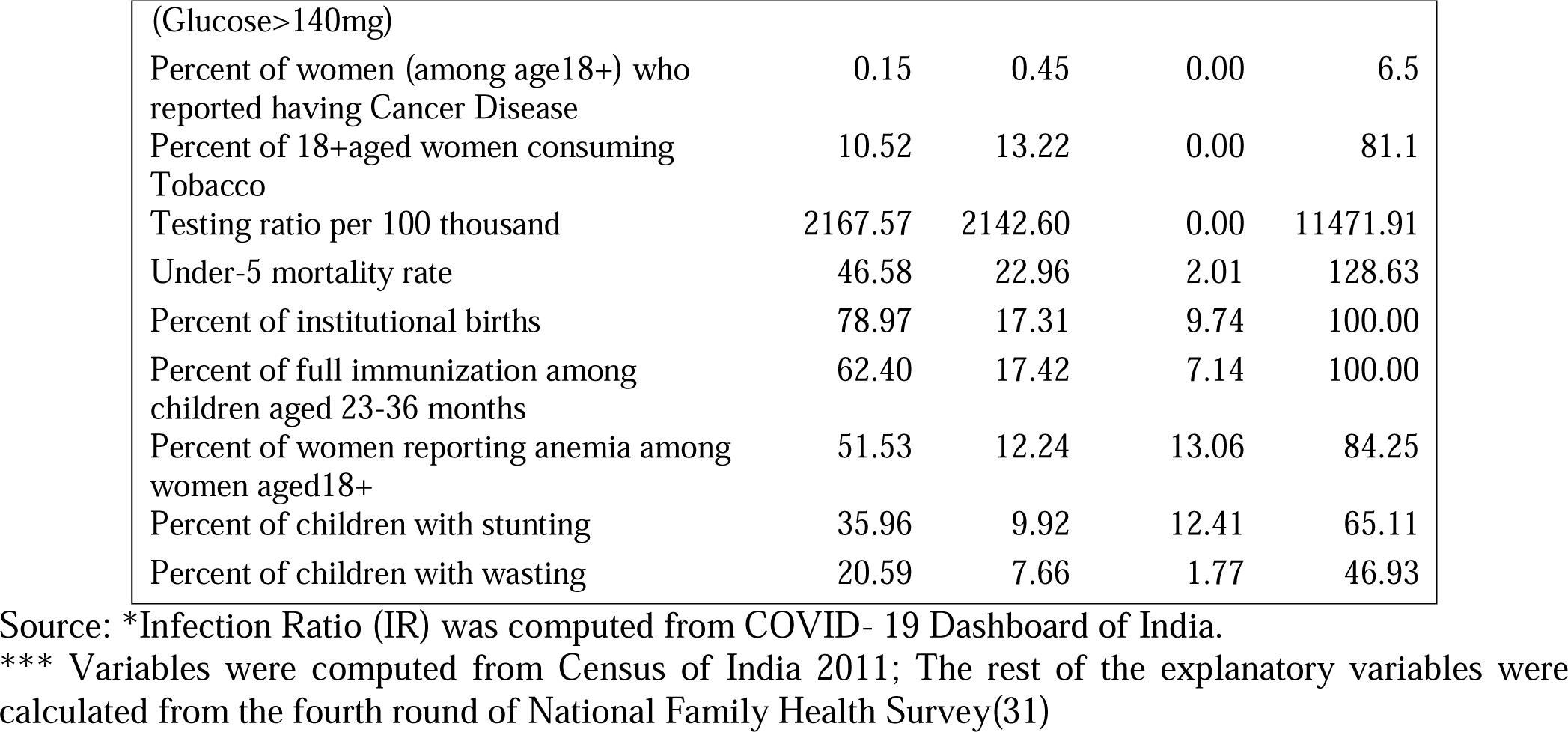
Summary statistics for the outcome and explanatory variables across 640 districts of India.

We also observed massive variation in the districts’ socio-economic variables, viz, percent Hindu, urban, and ST population vary between 0 percent to 100 percent in India’s districts. On average, 2.92 persons sleep in one room in the Indian districts. On average, 48.14 per cent of the households have water facilities, and 61.41 per cent have toilet facilities within the household premise. Hygiene practice of availability of soap ranges from less than 17.20 per cent to 98.90 per cent in India.

with response t to health-related variables as well, there exists a wide disparity. While the testing ratio ranged from 0 to11471 persons per 100 hundred thousand, the percentage of full immunization among children ranged from 7.14 to 100. The tobacco consumption among women ranged from 0.8 to 88 percent.

The regression analysis present in **Table 2** displays both unadjusted and adjusted coefficients of the exploratory variables. Among demographic variables, population density and district outmigration were associated with IRs in the unadjusted model. In contrast, several socio-economic variables, such as the literate population, ST population, and urban population, were significantly associated. The percentage of women whose husbands were away for six months is associated negatively with IR (β: −0.65, CI: −5.02-7.99). Compared to those with low literacy rates, districts with higher literacy rates showed 7.1 times higher likelihood of COVID-19 infection. Districts with higher levels of infrastructure reported higher levels of IRs. Among the health-related variables, percentage of women with diabetes (glucose>140), testing ratio and percentage of children immunized were significantly positively correlated with IR in the unadjusted model. Districts with high diabetic patients have 21.05-fold higher chance of COVID-19 infection in comparison to those with low prevalence of diabetes. However, percentage of women consuming tobacco and percentage of children with stunting condition was associated positively with COVID-19 prevalence.

**Table 2.** Regression analysis of district correlates and Infection ratio (IR) across 640 districts of India^1^.

| Models | Unadjusted model |  |  | Adjusted model |  |  |
| --- | --- | --- | --- | --- | --- | --- |
| | β-Coeff. | p-value | 95% CI | β-Coeff. | p-value | 95% CI |
| **Demographic variables** |  |  |  |  |  |  |
| **percent of 60 years and above population** | -1.42 | 0.84 | (-15.58, -12.74) | -7.28 | 0.5 | (-28.31, -13.75) |
| Percent of women whose husbands are away for the last 6months | -13.08** | 0 | (-18.15, -8.01) | 1.48 | 0.65 | (-5.02, -7.99) |
| Population density | 0.06** | 0 | (0.06, -0.07) | 0.05** | 0 | (0.04-0.06) |
| **Socio-economic variables** |  |  |  |  |  |  |
| Percent of literate population | 7.15** | 0 | (4.44, -9.85) | 2 | 0.4 | (-2.63, -6.62) |
| Percent of SC population | 0.43 | 0.79 | (-2.76, -3.62) | 3.92* | 0.04 | (0.12-7.72) |
| Percent of ST population | -1.22* | 0.03 | (-2.29, -0.14) | 0.32 | 0.7 | (-1.34, -1.99) |
| Percent of Hindu population | -0.09 | 0.87 | (-1.18, -1.0) | 0.29 | 0.71 | (-1.24, -1.83) |
| Percent of Urban population | 7.27** | 0 | (6.01, -8.53) | 3.08** | 0 | (1.05-5.11) |
| Avg persons sleeping in a room | -28.21 | 0.3 | (-81.04, -24.62) | -1.7 | 0.97 | (-79.56, -76.16) |
| **Infrastructure variables** |  |  |  |  |  |  |
| Percent of households with soap availability | 3.50** | 0 | (2.04, -4.96) | -0.03 | 0.98 | (-2.24, -2.19) |
| Percent of households with water availability within the premise | -0.17 | 0.75 | (-1.27, -0.92) | -0.53 | 0.37 | (-1.71, -0.64) |
| Percent of households with Toilet facility within the premise | 2.41** | 0 | (1.38, -3.45) | 0.42 | 0.7 | (-1.73, -2.57) |
| **Health-related variables** |  |  |  |  |  |  |
| Percent of women with Diabetes (Glucose>140mg) | 21.05** | 0 | (8.06, -34.05) | 3 | 0.66 | (-10.57, -16.58) |
| Percent of women (among age18+) who reported having Cancer Disease | 25.6 | 0.44 | (-39.49, -90.68) | 23.65 | 0.41 | (-32.26, -79.56) |
| Percent of 18+aged women consuming Tobacco | -2.68* | 0.02 | (-4.87, -0.49) | 0.34 | 0.8 | (-2.27, -2.96) |
| Testing ratio per 100 thousand | 0.03** | 0 | (0.02, -0.04) | 0.03** | 0 | (0.01-0.04) |
| Under-5 mortality rate | -2.59** | 0 | (-3.84, -1.33) | -1.16 | 0.14 | (-2.71, -0.38) |
| Percent of institutional births | 2.94** | 0 | (1.28, -4.6) | 0.13 | 0.92 | (-2.33, -2.59) |
| Percent of full immunization among children aged 23-36 months | 0.81 | 0.34 | (-0.86, -2.48) | -1.26 | 0.17 | (-3.07, -0.55) |
| Percent of women reporting anemia among women aged18+ | 0.39 | 0.75 | (-1.98, -2.77) | 0.98 | 0.79 | (-1.46,3.42) |
| Percent of children with stunting | -4.58** | 0 | (-7.49, -1.67) | 3.05 | 0.18 | (-1.37, -7.47) |

<sup>1</sup>All variables are computed at the district level

|  |  |  |  |  |  |  |
| --- | --- | --- | --- | --- | --- | --- |
| Percent of children with wasting | -1.17 | 0.55 | (-4.97, -2.63) | 2.02 | 0.35 | (-2.19, -6.23) |

\*p < 0.05, \*\*p < 0.01
Source: Author's Computation

In the adjusted model, the association between IRs and most of the correlates becomes statistically insignificant. After controlling the roles of demographic, socio-economic, infrastructural, and health-related variables in the adjusted model, district-level population density (β: 0.05, CI:004-0.06), the percentage of urban population (β:3.08, CI: 1.05-5.11), percentage of Scheduled Caste Population (β: 3.92, CI: 0.12-7.72) and district level testing ratio (β: 0.03, CI: 0.01-0.04) were positively associated with the prevalence of COVID-19.

## Summary and Discussion

In terms of the total number of positive cases, India ranked third after the US and Brazil, reporting more than one million COVID-19 cases as on July 31, 2020. A further concern is India’sCOVID-19 curve remaining on the upward trajectory with no sign of bending like Italy and the UK. Due to different demographic transition phases in the states, the trajectory of COVID-19 and related intervention cannot be uniform. Our result illustrated the differences in COVID-19 cases at the state and district levels with few critical findings.

First, the spread of COVID-19 has been increasing over time. The average bi-weekly cases show that the new, infected, recovered, and deceased cases are growing nationally. Besides, the 80 per cent of the new patients, and 90 per cent of the deaths are concentrated in nine Indian states (Maharashtra, Delhi, Tamil Nadu, Karnataka, Andhra Pradesh, Telangana, West Bengal, Gujarat, and Uttar Pradesh). On July 31,2020, IR in India is 42.38 per 100 hundred thousand population, with the highest in Andhra Pradesh (145.0) followed by Maharashtra (123.6) and the lowest in Chhattisgarh (10.1). Only two Union Territories (Lakshadweep and Daman & Diu) have zero IR. The most affected cities are Mumbai and Pune in Maharashtra, Kolkata in West Bengal, Bangalore in Karnataka, and Chennai in Tamil Nadu, Ahmadabad in Gujarat, and Gurgaon and Noida in NCR (National Capital Region). The study identifies the districts at higher risk of coronavirus infection in the southern, northern, and western states. We also found six high-risk COVID-19infected cities (Mumbai, Chennai, Thane, Pune, Bengaluru, and Hyderabad) in India. The apparent concern is that these states also contribute significantly to the Indian economy(32). Another important observation of this study is that districts bordering the six metropolitan cities were observed to be India’s highest hot spots, possibly because they contribute the largest share of migrants and commuters to these megacities. This study indicates, in addition to Maharashtra, Delhi, Tamil Nadu, and Gujarat, states with low socio-economic indicators such as the EAG states (Bihar, Chhattisgarh, Jharkhand, Madhya Pradesh, Uttar Pradesh, Rajasthan, Odisha, Uttarakhand) are more affected due to the lack of health care facilities.

Secondly, this study examined the district level correlates with the COVID-19 infection ratio in India. Our research reveals that the district’s infection ratio of COVID-19 is associated with various socio-economic variables. However, we observed a statistically significant association only with a limited variable. After adjusting the role of socio-economic and health-related factors, the COVID-19 infection rate was found to be higher in the districts with a higher level of population density, a higher percentage of the urban population, a higher percentage of deprived castes and a higher level of testing ratio. High population density might lead to social distancing challenges, thereby districts with higher density have higher infection ratio.

Similarly, as the percentage of the urban population increases, the chances of unavoidable economic activities might increase, which exposes more people to the Coronavirus. Previous studies also showed that higher population densities in congested slum areas and large towns accelerated COVID-19 infection and mortality rates(33–35). The congestion, slum concentrations, inadequate housing, and sanitation in poor urban areas may explain such high disease. A positive association between COVID-19, IRs and testing ratio indicates underreporting of COVID-19, in districts where the testing ratio is low.

Studies based on individual data show that older people are more vulnerable to COVID-19 infections (36,37). This study also identified that pre-existing diabetes is positively associated with COVID-19 disease(36,37). In our research, we didn’t find such associations, possibly because of the study design. Unlike these studies, we are identifying macro-level correlates of the COVID-19 infection rate. Interestingly, as population belonging to deprived castes such as SCs increase, the chances of COVID-19 increase since SCs are more vulnerable and daily laborer they may have higher infection chances.

## Conclusion

The COVID-19 pandemic is expected to have a long-term impact on health, economy, and social processes globally, including India. Only a clear understanding of the disease’s spatial distribution and its determinants will help to formulate policies and interventions. Therefore, the possible risk factors should be included in policy preparedness and implementation during the COVID-19 pandemic.

We found that population density, urban residence, Scheduled Caste population, and testing rates are significantly correlated with the infection ratio (IR). As in urban areas, the population density is very high, and social distancing is challenging to maintain, the role of government is crucial in combating the pandemic. By ensuring the health and hygiene-related facilities, (providing adequate clean water, adequate sanitation, and sewerage facilities, cleaning the city, maintaining quarantine centers and public health care institutions, etc.), and improving public distribution system to ensure minimum food supply, especially among the urban poor and other deprived sub-groups, can help to control the spread of COVID-19 infection.

More tests are required to classify patients with asymptomatic conditions. Currently, India has a population of over 1.3 billion, but till July 31, approximately 18.8 million (lesser than 2 percent) tests have been carried out. Simultaneously, people’s negligent behavior towards COVID-19 protocols (say not following the social distancing norms, not wearing the mask in pubic place, and coughing without covering mouth) put them at a higher risk. Finally, there is need to improve infrastructure (hospitals, ventilators, PPE kits), and human resources (doctors, nurses, and frontline workers) in healthcare facilities. Our analysis does have a few limitations. First, there is a possibility of under-reporting positive and fatal cases due to a lack of testing or social stigma. Hence our data gives the most conservative estimates of infection ratio. Second, for most cases, the patients’ level of information (such as age, sex, and co-morbidity) is unavailable. Therefore, we analyzed the district level determinants instead of individual-level determinants. Thus, our results identified the major correlates only at the district level. Finally, we analyzed the number of confirmed cases rather than the number of active cases, as the later considers the recovery rate and reflects the health service available in a region. We used the number of confirmed cases as the primary indicator of the spread of the infection. Despite these limitations, the study’s merit lies in bringing together spatial-demographic vulnerabilities prevalent across the nation during the pandemic period. To sum up, the study findings identified the district level indication of COVID-19 and their demographic and socio-economic features.

## Data Availability

The data is publicly available.

https://www.covid19india.org/

## Supporting Information

**S1 Table.**
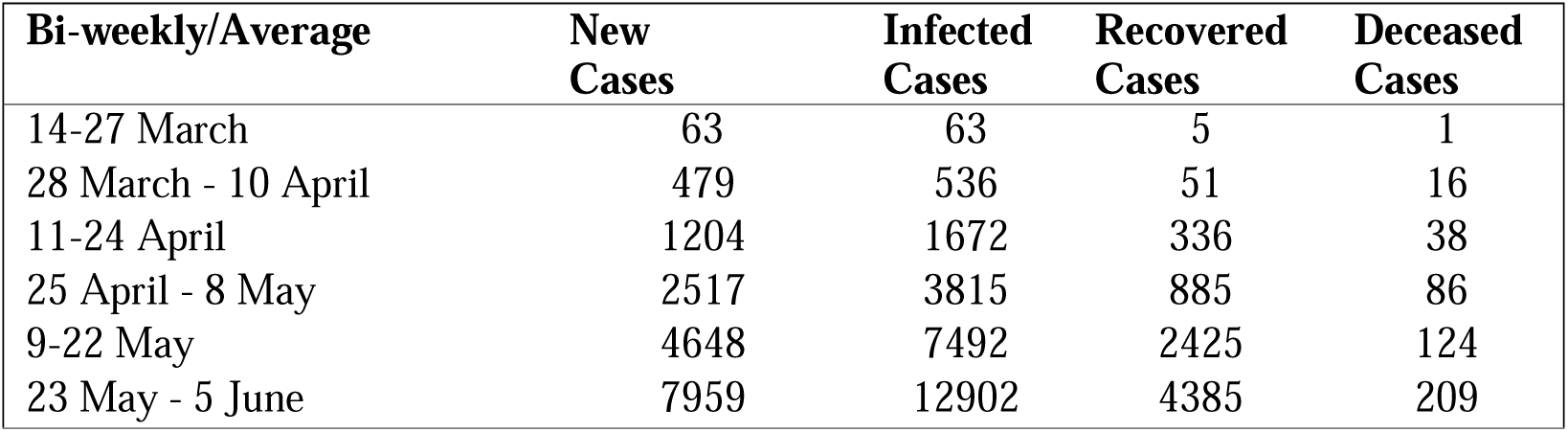
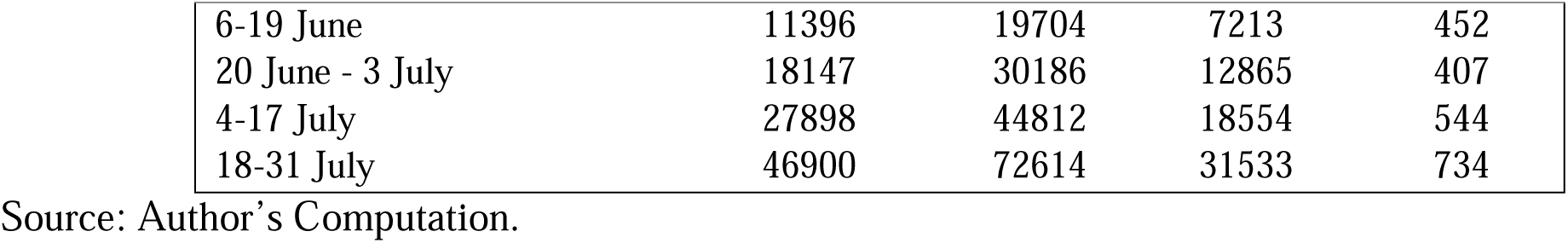
Average number of new cases, recovered, infected, deceased cases bi-weekly (14 days) in India (March 14, 2020 – July 31, 2020)

| Bi-weekly/Average | New Cases | Infected Cases | Recovered Cases | Deceased Cases |
| --- | --- | --- | --- | --- |
| 14-27 March | 63 | 63 | 5 | 1 |
| 28 March - 10 April | 479 | 536 | 51 | 16 |
| 11-24 April | 1204 | 1672 | 336 | 38 |
| 25 April - 8 May | 2517 | 3815 | 885 | 86 |
| 9-22 May | 4648 | 7492 | 2425 | 124 |
| 23 May - 5 June | 7959 | 12902 | 4385 | 209 |
| 6-19 June | 11396 | 19704 | 7213 | 452 |
| 20 June - 3 July | 18147 | 30186 | 12865 | 407 |
| 4-17 July | 27898 | 44812 | 18554 | 544 |
| 18-31 July | 46900 | 72614 | 31533 | 734 |

Source: Author's Computation.

**S2 Table.**
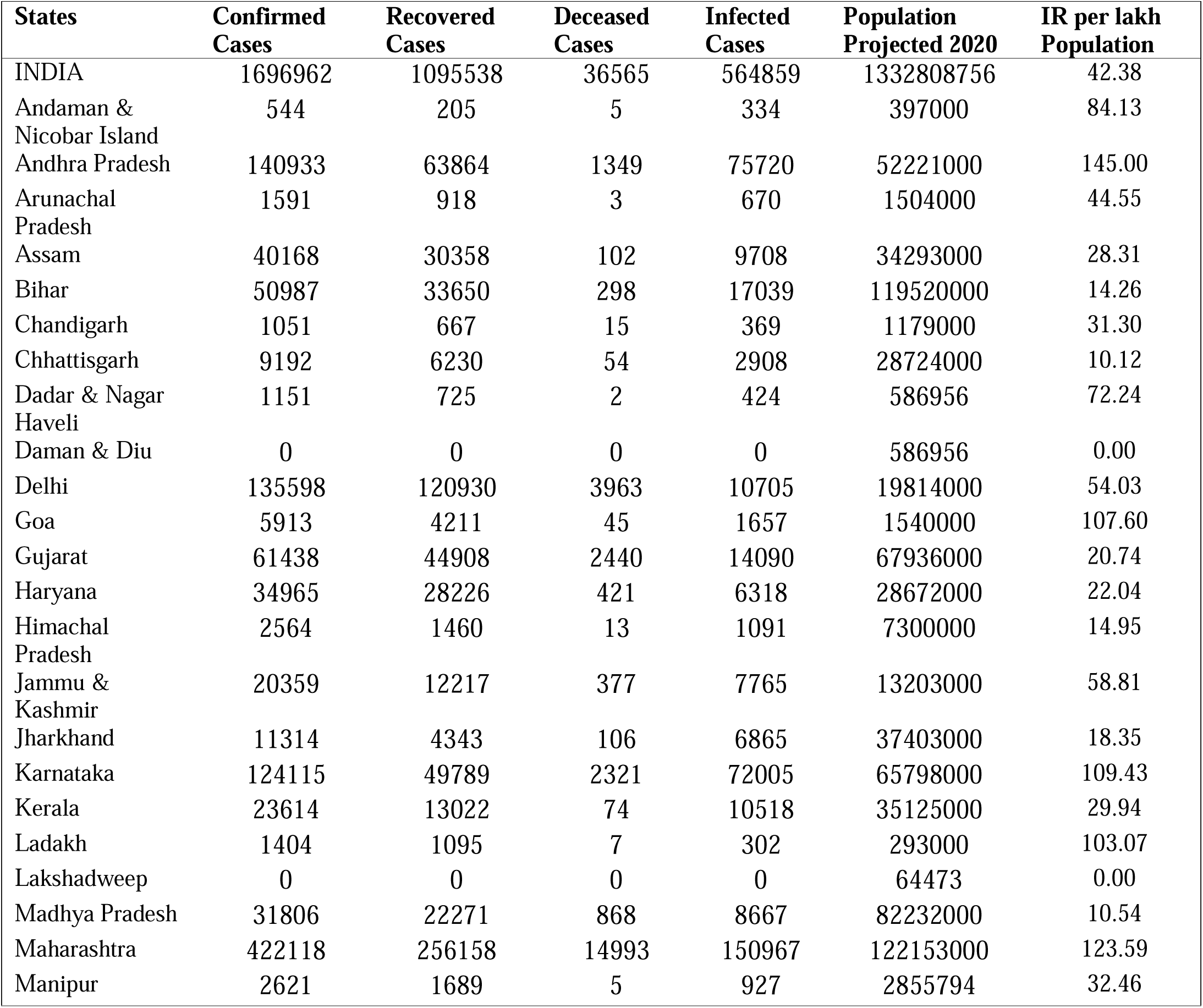
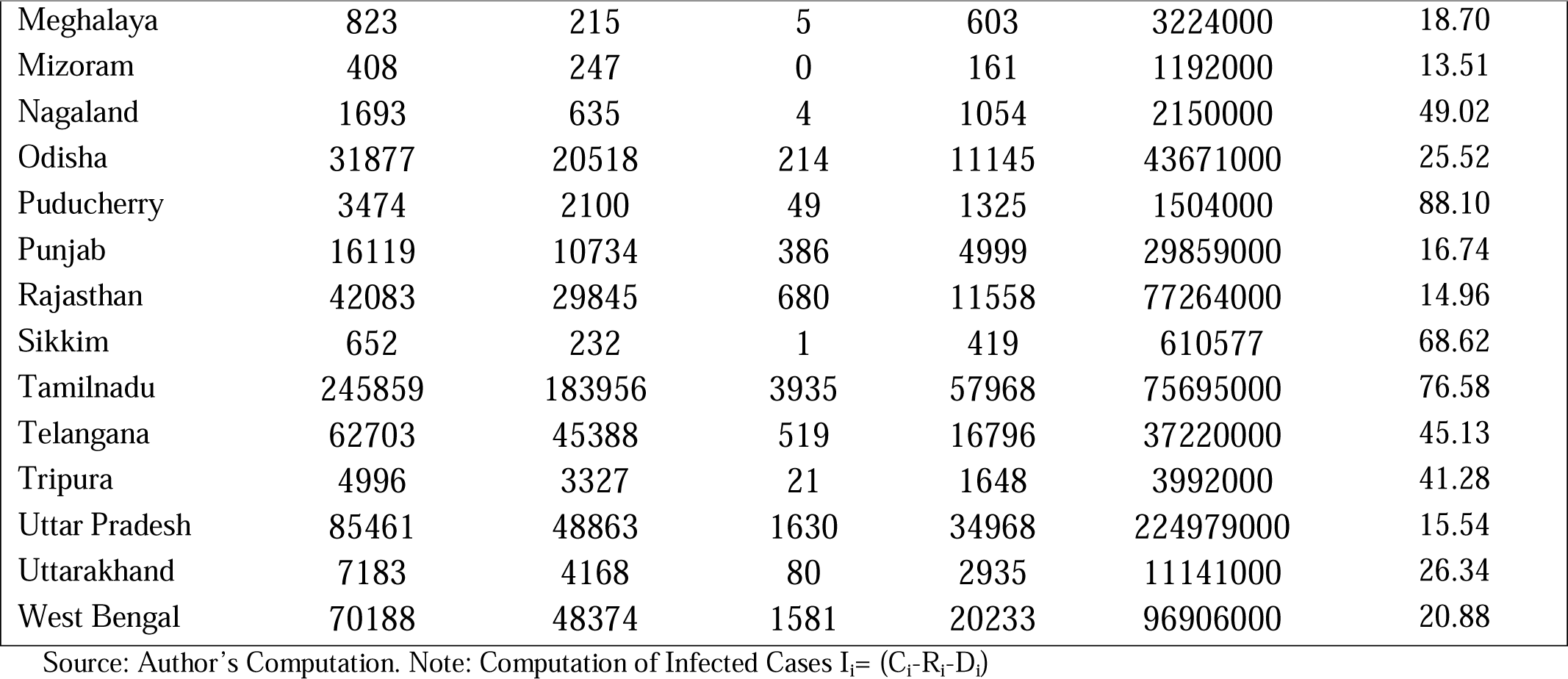
State-wise Infection Ratio (IR) in India (till July 31, 2020)

**S3 Table.**
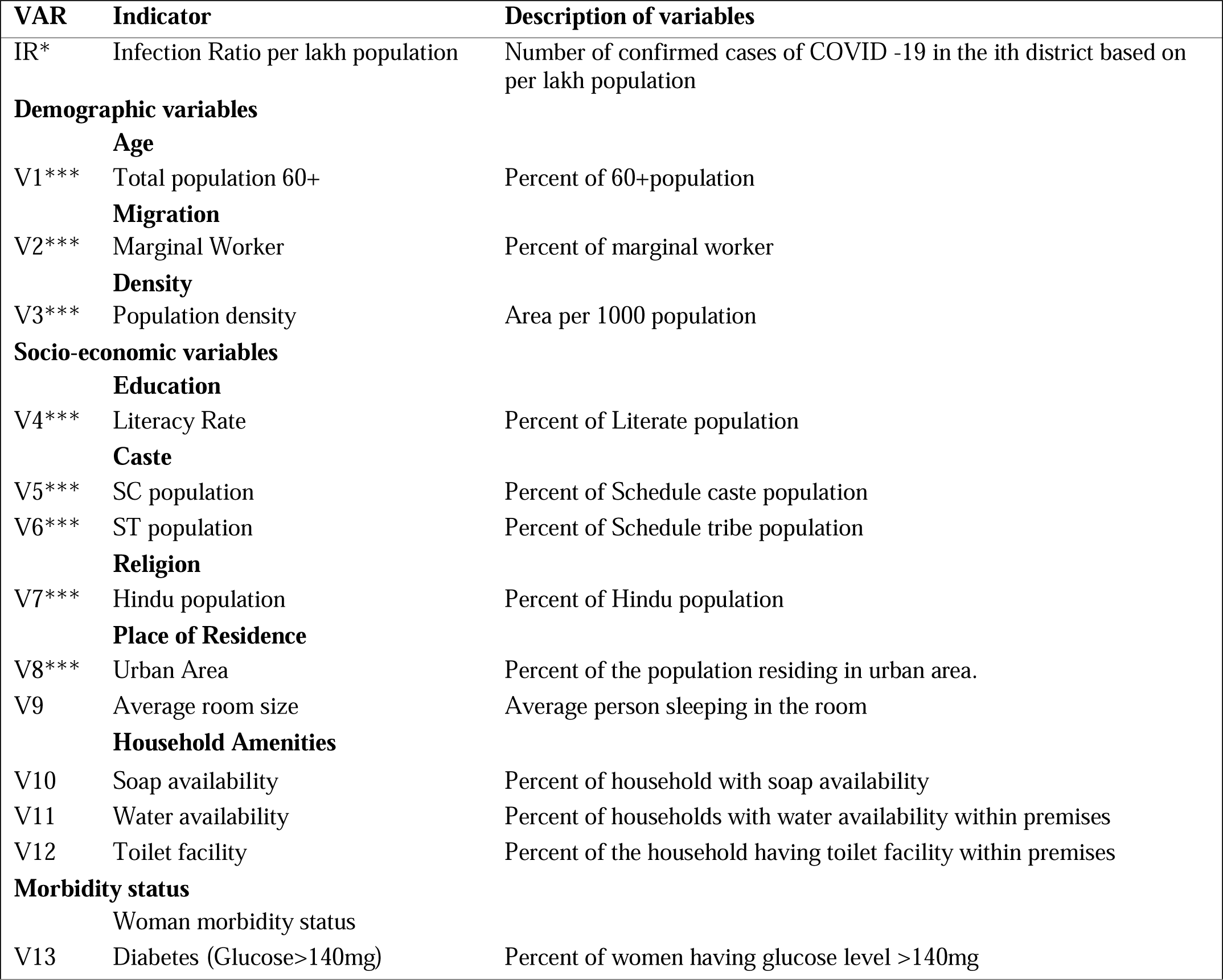
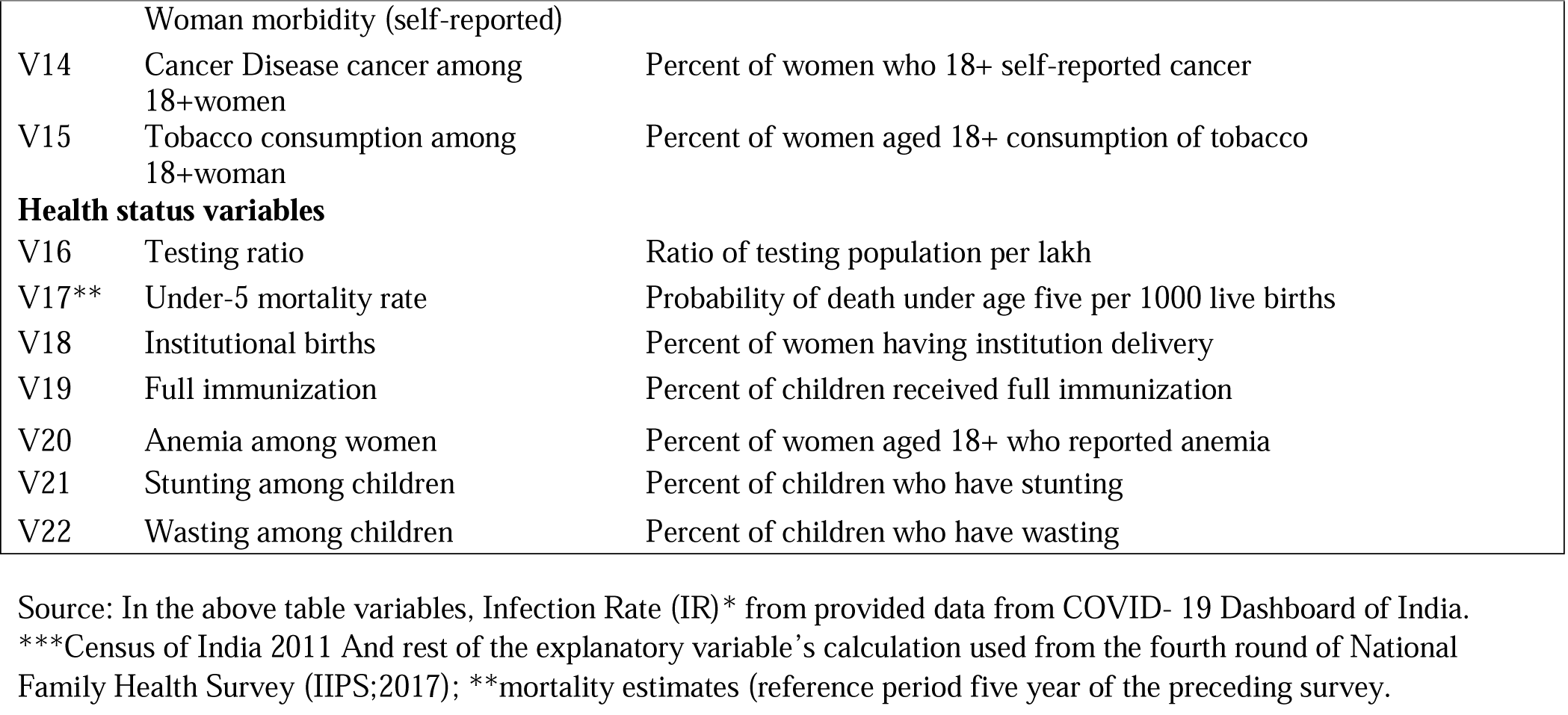
Brief description of each indicator used in this analysis.

## References

1. Worldometers. COVID-19 coronavirus pandemic. Retrieved June 30, 2020, from https://www.worldometers.info/coronavirus/. 2020.

2. Dwivedi LK, Rai B, Shukla A, Dey T, Ram U. Assessing the Impact of Complete Lockdown on COVID-19 Infections in India and its Burden on Public Health Facilities A Situational Analysis Paper for Policy Makers International Institute for Population Sciences, Mumbai. 2020.

3. Schueller E, Klein E, Tseng K, Kapoor G, Joshi J, Sriram A, et al. COVID-19 in India: Potential Impact of the Lockdown and Other Longer-Term Policies. Cent Dis Dyn Econ Policy. 2020;

4. Wang T, Du Z, Zhu F, Cao Z, An Y, Gao Y, et al. Comorbidities and multi-organ injuries in the treatment of COVID-19. Lancet. 2020;395(10228):e52.

5. Rao HR, Vemprala N, Akello P, Valecha R. Retweets of officials’ alarming vs reassuring messages during the COVID-19 pandemic: Implications for crisis management. Int J Inf Manage. 2020;(June):102187.

6. Charlton E. This is how India is reacting to the coronavirus pandemic. World Economic Forum. 2020;

7. Soni P. These are the coronavirus quarantine facilities in India. Bus Insid India. 2020;March 25,.

8. Philip Debraj Ray Subramanian MS, Philip M, Ray D, Subramanian S, Parade Road St Thomas N, Chennai M. NBER WORKING PAPER SERIES DECODING INDIA’S LOW COVID-19 CASE FATALITY RATE Decoding India’s Low Covid-19 Case Fatality Rate †. 2020.

9. Dowd JB, Andriano L, Brazel DM, Rotondi V, Block P, Ding X, et al. Demographic science aids in understanding the spread and fatality rates of COVID-19. Proc Natl Acad Sci U S A. 2020;117(18):9696–8.

10. Nepomuceno MR, Acosta E, Alburez-Gutierrez D, Aburto JM, Gagnon A, Turra CM. Besides population age structure, health and other demographic factors can contribute to understanding the COVID-19 burden. Proc Natl Acad Sci [Internet]. 2020;117(25):13881–3. Available from: https://www.pnas.org/content/117/25/13881

11. Mahajan P, Kaushal J. Epidemic Trend of COVID-19 Transmission in India During Lockdown-1 Phase. J Community Health. 2020;(0123456789).

12. Srivastava A, Tamrakar V, Naaz Akhtar S, Kumar K, Chand Saini T, Saikia N. Geographical Variation in COVID-19 Cases, Prevalence, Recovery and Fatality Rate by Phase of National Lockdown in India, March 14-May 29, 2020. 2020;

13. MHA. New Guidelines on the measures to be taken by Ministries/Department of Government of India, State/UT Governments and State/UT authorities for containment of COVI-19 in the country for the extended period of National Lockdown for a further period of two we [Internet]. 40-3/2020-DM-I (A) India; 2020. Available from: 164.100.117.97/WriteReadData/userfiles/MHA Order Dt. 1.5.2020 to extend Lockdown period for 2 weeks w.e.f. 4.5.2020 with new guidelines.pdf

14. Singh SS. Corona Virus.The mystery of the low COVID-19 numbers in West Bengal. The Hindu. 2020 May;

15. Acharya R, Porwal A. A vulnerability index for the management of and response to the COVID-19 epidemic in India: an ecological study. Lancet Glob Heal [Internet]. 2020 Aug 16; Available from: https://doi.org/10.1016/S2214-109X(20)30300-4

16. Golechha M, Panigrahy RK. COVID-19 and heatwaves: a double whammy for Indian cities. Lancet Planet Heal. 2020;4:e315–6.

17. COVID-19-India. COVID-19 India Dashboard. 2020.

18. MOHFW. COVID-19 INDIA, Ministry of Health and Family Welfare, Government of India. 2020.

19. IIPS. International Institute for Population Science India: National Family Health Survey (NFHS-4), 2015–16: India. Mumbai; 2017.

20. ORGI. Office of the Registrar General & Census Commissioner, India (ORGI), Provisional Population TotalsPaper 2 of India & States/UTs. 2011.

21. Saikia N, Bora JK, Jasilionis D, Shkolnikov VM. Disability Divides in India: Evidence from the 2011 Census. PLoS One [Internet]. 2016 Aug 4;11(8):e0159809. Available from: https://doi.org/10.1371/journal.pone.0159809

22. Striessnig E, Bora JK. Under-Five Child Growth and Nutrition Status: Spatial Clustering of Indian Districts. Spat Demogr. 2020;8(1):63–84.

23. Guilmoto CZ, Saikia N, Tamrakar V, Bora JK. Excess under-5 female mortality across India: a spatial analysis using 2011 census data. Lancet Glob Heal. 2018 Jun;6(6):e650–8.

24. Nandi A, Balasubramanian R, Laxminarayan R. Who is at the highest risk from COVID-19 in India? Analysis of health, healthcare access, and socioeconomic indicators at the district level. medRxiv [Internet]. 2020 Jan 1;2020.04.25.20079749. Available from: http://medrxiv.org/content/early/2020/06/09/2020.04.25.20079749.abstract

25. Watkins SC, Menken JA, Bongaarts J. Demographic Foundations of Family Change. Am Sociol Rev [Internet]. 1987;52(3):346–58. Available from: http://www.jstor.org/stable/2095354

26. Yi Z. Changing Demographic Characteristics and the Family Status of Chinese Women. Popul Stud (NY) [Internet]. 1988 Jul 1;42(2):183–203. Available from: https://doi.org/10.1080/0032472031000143316

27. Zeng Y, Vaupel J, Wang Z. Household projection using conventional demographic data. Popul Dev Rev. 1998;24:59.

28. Saikia N, Bora JK, Jasilionis D, Shkolnikov VM. Disability divides in India: Evidence from the 2011 census. PLoS One. 2016;11(8):1–13.

29. Moran PAP. Notes on Continuous Stochastic Phenomena. Biometrika [Internet]. 1950 May 30;37(1/2):17–23. Available from: http://www.jstor.org/stable/2332142

30. Weinreb A, Gerland P, Fleming P. Hotspots and Coldspots: Household and village-level variation in orphanhood prevalence in rural Malawi. Demogr Res [Internet]. 2008 Jul 15;19(32):1217. Available from: https://pubmed.ncbi.nlm.nih.gov/20148129

31. IIPS. International Institute for Population Science India: National Family Health Survey (NFHS-4), 2015–16: India [Internet]. Mumbai; 2017. Available from: http://rchiips.org/nfhs/NFHS-4Reports/India.pdf

32. Singh M, Neog Y. Contagion effect of COVIDLJ19 outbreak: Another recipe for disaster on Indian economy. J Public Aff. 2020 May 1;

33. You H, Wu X, Guo X. Distribution of COVID-19 Morbidity Rate in Association with Social and Economic Factors in Wuhan, China: Implications for Urban Development. Int J Environ Res Public Health [Internet]. 2020 May 14;17(10):3417. Available from: https://pubmed.ncbi.nlm.nih.gov/32422948

34. Hamidi S, Sabouri S, Ewing R. Does Density Aggravate the COVID-19 pandemic? J Am Plan Assoc [Internet]. 2020 Jun 18;1–15. Available from: https://doi.org/10.1080/01944363.2020.1777891

35. Li R, Richmond P, Roehner BM. Effect of population density on epidemics. Phys A Stat Mech its Appl [Internet]. 2018;510:713–24. Available from: http://www.sciencedirect.com/science/article/pii/S0378437118308896

36. Richardson S, Hirsch JS, Narasimhan M, Crawford JM, McGinn T, Davidson KW, et al. Presenting Characteristics, Comorbidities, and Outcomes Among 5700 Patients Hospitalized With COVID-19 in the New York City Area. JAMA [Internet]. 2020 Apr 22;323(20):2052–9. Available from: https://pubmed.ncbi.nlm.nih.gov/32320003

37. Zhou F, Yu T, Du R, Fan G, Liu Y, Liu Z, et al. Clinical course and risk factors for mortality of adult inpatients with COVID-19 in Wuhan, China: a retrospective cohort study. Lancet [Internet]. 2020 Mar 28;395(10229):1054–62. Available from: https://doi.org/10.1016/S0140-6736(20)30566-3

